# Clinical Characteristics of Hospitalized Covid-19 Patients in New York City

**DOI:** 10.1101/2020.04.19.20062117

**Authors:** Ishan Paranjpe, Adam J Russak, Jessica K De Freitas, Anuradha Lala, Riccardo Miotto, Akhil Vaid, Kipp W. Johnson, Matteo Danieletto, Eddye Golden, Dara Meyer, Manbir Singh, Sulaiman Somani, Sayan Manna, Udit Nangia, Arjun Kapoor, Ross O’Hagan, Paul F O’Reilly, Laura M Huckins, Patricia Glowe, Arash Kia, Prem Timsina, Robert M Freeman, Matthew A Levin, Jeffrey Jhang, Adolfo Firpo, Patricia Kovatch, Joseph Finkelstein, Judith A Aberg, Emilia Bagiella, Carol R Horowitz, Barbara Murphy, Zahi A Fayad, Jagat Narula, Eric J Nestler, Valentin Fuster, Carlos Cordon-Cardo, Dennis S Charney, David L Reich, Allan C Just, Erwin P Bottinger, Alexander W Charney, Benjamin S Glicksberg, Girish N Nadkarni, on behalf of the Mount Sinai Covid Informatics Center (MSCIC)

**Affiliations:** The Hasso Plattner Institute for Digital Health at the Mount Sinai; Department of Internal Medicine; Department of Genetics and Genomic Sciences; The Zena and Michael A. Wiener Cardiovascular Institute; Department of Population Health Science and Policy; The Pamela Sklar Division of Psychiatric Genomics; The Department of Psychiatry; Institute for Healthcare Delivery Science; Department of Pathology, Molecular and Cell-Based Medicine; Icahn Institute for Data Science and Genomic Technology; Mount Sinai Data Warehouse; Division of Infectious Diseases; Department of Medicine; The Charles Bronfman Institute for Personalized Medicine; The BioMedical Engineering and Imaging Institute; Department of Radiology; Department of Cardiology; The Nash Family Department of Neuroscience; The Friedman Brain Institute; The Office of the Dean; Department of Anesthesiology, Perioperative and Pain Medicine; Department of Environmental Medicine and Public Health; Institute for Exposomic Research; The Charles Bronfman Institute for Personalized Medicine, Icahn School of Medicine at Mount Sinai

**Author notes:** **Correspondence:** Please address correspondence to: Erwin P Bottinger, MD, Alexander W. Charney, MD, PhD, Benjamin S. Glicksberg, PhD, Girish N. Nadkarni, MD, MPH. These authors contributed equally (Ishan Paranjpe, Adam J Russak, Jessica K De Freitas, Anuradha Lala). These authors jointly supervised this work (Erwin P Bottinger, Alexander W Charney, Benjamin S Glicksberg, Girish N Nadkarni).

## Abstract

**Background:** The coronavirus 2019 (Covid-19) pandemic is a global public health crisis, with over 1.6 million cases and 95,000 deaths worldwide. Data are needed regarding the clinical course of hospitalized patients, particularly in the United States.

**Methods:** Demographic, clinical, and outcomes data for patients admitted to five Mount Sinai Health System hospitals with confirmed Covid-19 between February 27 and April 2, 2020 were identified through institutional electronic health records. We conducted a descriptive study of patients who had in-hospital mortality or were discharged alive.

**Results:** A total of 2,199 patients with Covid-19 were hospitalized during the study period. As of April 2^nd^, 1,121 (51%) patients remained hospitalized, and 1,078 (49%) completed their hospital course. Of the latter, the overall mortality was 29%, and 36% required intensive care. The median age was 65 years overall and 75 years in those who died. Pre-existing conditions were present in 65% of those who died and 46% of those discharged. In those who died, the admission median lymphocyte percentage was 11.7%, D-dimer was 2.4 ug/ml, C-reactive protein was 162 mg/L, and procalcitonin was 0.44 ng/mL. In those discharged, the admission median lymphocyte percentage was 16.6%, D-dimer was 0.93 ug/ml, C-reactive protein was 79 mg/L, and procalcitonin was 0.09 ng/mL.

**Conclusions:** This is the largest and most diverse case series of hospitalized patients with Covid-19 in the United States to date. Requirement of intensive care and mortality were high. Patients who died typically had pre-existing conditions and severe perturbations in inflammatory markers.

## INTRODUCTION

The Coronavirus disease (Covid-19) pandemic, caused by severe acute respiratory syndrome coronavirus-2 (SARS-CoV-2), has held the world at a standstill with its virulence. As of April 8, 2020, over 1.6 million people have been affected, and more than 95,000 patients have died worldwide^1^. In addition to being highly contagious, the disease manifestations and clinical course are variable, spanning from asymptomatic status to severe acute respiratory distress syndrome with multiorgan failure and death^2^.

Reports from China and Italy provided early data on disease presentation and management^3,4^ but also revealed varying geographic disease expressions. Over one-fourth of the world’s cases are now in the United States (US), with more than a third of all US cases ascertained in New York state alone, making it the current epicenter of the Covid-19 pandemic^5^.

As the number of cases continues to climb, hospitals are being stretched well-beyond capacity while facing challenges of insufficient personal protective equipment (PPE), ventilators, and workforce. Thus, understanding the clinical course of hospitalized Covid-19 patients is critical not only for providing optimal patient care but also to inform resource management in other locations across the US likely to experience similar case surges^6^.

The Mount Sinai Healthcare System (MSHS) is the largest academic health system in New York City and serves as an ideal platform to better understand the evolving landscape of Covid-19 across a diverse population. Here, we present the largest case series of patients hospitalized with laboratory confirmed Covid-19 to date in the US.

## METHODS

### Study Population

The MSHS serves a large, racially and ethnically diverse patient population. In this study, patient data came from 5 major hospitals: the Mount Sinai Hospital located in East Harlem, Manhattan; Mount Sinai Morningside located in Morningside Heights, Manhattan; Mount Sinai West located in Midtown and the West Side, Manhattan; Mount Sinai Brooklyn located in Midwood, Brooklyn; and Mount Sinai Queens located in Astoria, Queens. We included patients who were at least 18 years old, had a laboratory-confirmed Covid-19 infection, and were admitted to any of the aforementioned 5 MSHS hospitals between February 27 and 12 a.m. April 2, 2020 (time of data freeze). A confirmed case of Covid-19 was defined by a positive reverse transcriptase polymerase chain reaction (RT-PCR) assay of a specimen collected via nasopharyngeal swab. The Mount Sinai Institutional Review Board approved this research under a broad regulatory protocol allowing for analysis of limited patient-level data.

### Data Collection

The dataset was obtained from different sources and aggregated by the New York Covid Informatics Taskforce (NYCIT) (Further description of NYCIT is provided in Supplementary Material). We obtained demographics, diagnosis codes (International Classification of Diseases-9/10-Clinical Modification (ICD-9/10-CM) codes and procedures, as well as vital signs and laboratory measurements during hospitalization. Demographics included age, sex, and language, as well as race and ethnicity in the electronic health records. Racial groups included White, Black or African American, Asian, Pacific Islander, Other, and Unknown. Ethnic groups included Non-Hispanic/Latino, Hispanic/Latino, or Unknown. All vital signs and laboratory values were obtained as part of indicated clinical care.

### Definitions of Pre-existing Conditions

We defined a pre-existing condition as the presence of diagnosis codes associated with specific diseases. Diagnoses and corresponding ICD codes are provided in **Supplemental Table 1**.

### Definitions of Outcomes

We assessed in-hospital mortality and admission to intensive care.

### Statistical Analysis

We used descriptive statistics were used to summarize the data; results are reported as medians and interquartile ranges or means and standard deviations, as appropriate. Categorical variables were summarized as counts and percentages. We visualized length of stay (LOS) using a cumulative incidence function with competing risks for mutually exclusive events of in-hospital mortality or discharge. Patients who were still hospitalized at the time of data freeze were regarded as having a censored LOS. We assumed censored observations from patients with ongoing hospitalization will not exceed the longest LOS in our dataset when calculating the restricted mean LOS. No imputation was made for missing data. Analysis was performed with R^7^.

## RESULTS

A consort diagram of included patients and outcomes is depicted in **Supplementary Figure 1**.

### Demographic and Clinical Characteristics

From February 27 to April 2, 2020, 2,199 Covid-19 positive patients were hospitalized at one of five MSHS New York City hospitals. At the time of writing this report, 1,121 (51%) patients remained hospitalized and 1,078 (49%) completed their hospital course, with 768 discharges and 310 deaths. **Figure 1** details the number of patients admitted to the hospital per day and the total number of patients admitted cumulatively over time. During the study period, the trend of hospital admissions per day consistently increased.

**Figure 1:**
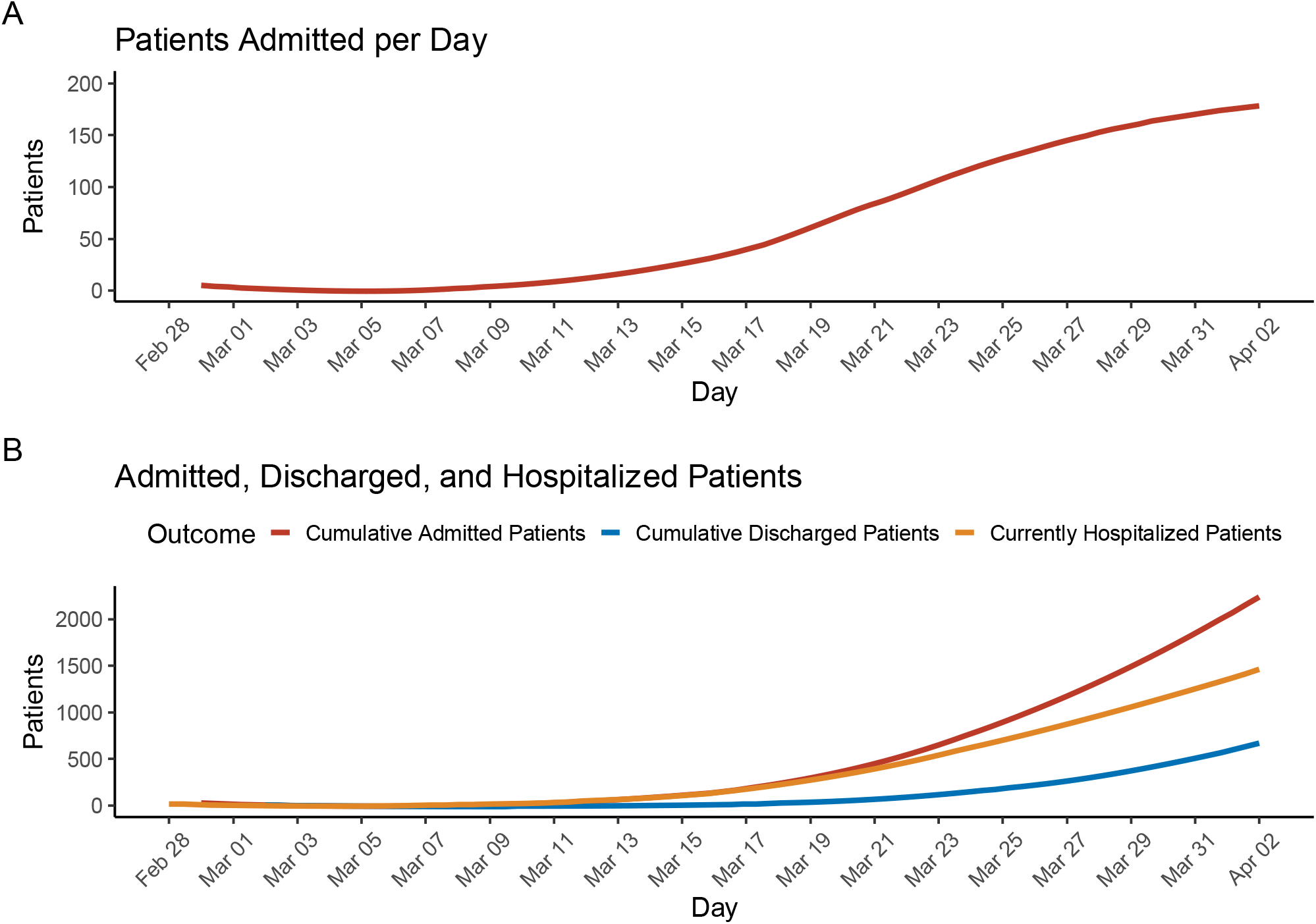
Hospital Admissions of Covid-19 patients within our cohort. <u>Panel A</u>: The number of total patients (n = 2,199) admitted each day to one of the hospitals for the duration of the study period. <u>Panel B</u>: The number of patients cumulatively admitted, cumulatively discharged, or still hospitalized by day.

Patient demographics, pre-existing conditions as well as vital signs and laboratory values at the time of admission are displayed in Table 1. Median age was 65 years with only 3% of patients less than 30 years and 36% over 70 years. The proportion of men was higher (59%) than women (41%) and 25% had their race identified as White, 25% as African American, and 3% as Asian. One quarter of the population has their ethnicity identified as Hispanic/Latino. More than half of the population had at least one pre-existing condition. Specifically, 37% presented with a history of hypertension, 27% with diabetes mellitus, 16% with coronary artery disease, 10% with heart failure and 9% with chronic kidney disease.

**Table 1.** Characteristics of Hospitalized Covid-19 Patients at Baseline (n=2,199)

| Characteristics of Admission |  | N with characteristic available |
| --- | --- | --- |
| Admission Source, n (%) |  |  |
| Emergency Department | 1558 (71) | 2199 |
| Other | 641 (29) |  |
| Race, n (%) |  |  |
| White | 554 (25.2) | 2199 |
| Black or African American | 543 (24.7) |  |
| Asian | 74 (3.4) |  |
| Pacific Islander | 25 (1.1) |  |
| Other | 912 (41.5) |  |
| Unknown | 91 (4.1) |  |
| Ethnicity, n (%) |  |  |
| Hispanic/Latino | 576 (26.2) | 2199 |
| Non-Hispanic/Latino | 1305 (59.4) |  |
| Unknown | 318 (14.5) |  |
| Age, Median (IQR) | 65 (54-76) | 2199 |
| Age Groups, n (%) |  |  |
| 18-30 | 73 (3.3) | 2199 |
| 31-40 | 179 (8.1) |  |
| 41-50 | 225 (10.2) |  |
| 51-60 | 395 (18.0) |  |
| 61-70 | 527 (24.0) |  |
| 71-80 | 444(20.2) |  |
| 81-90 | 274 (12.5) |  |
| 91 or older | 82 (3.7) |  |
| Sex, n (%) |  |  |
| Male | 1293 (58.8) | 2199 |
| Female | 906 (41.2) |  |
| Body Mass Index in kg/m<sup>2</sup>, Median (IQR) | 28 (6) | 1316 |
| Previous Medical History, n (%) |  |  |
| Atrial Fibrillation | 156 (7.1) | 2199 |
| Asthma | 180 (8.2) |  |
| Coronary Artery Disease | 343 (15.6) |  |
| Cancer | 151 (6.9) |  |
| Chronic Kidney Disease | 207 (9.4) |  |
| Chronic Obstructive Pulmonary Disease | 113 (5.1) |  |
| Diabetes Mellitus | 583 (26.5) |  |
| Heart Failure | 217 (9.9) |  |
| Hypertension | 812 (37) |  |
| Stroke | 153 (7) |  |
| Vital Signs at Hospital Admission |  |  |
| Heart Rate in beats per min, Median (IQR) | 95 (83-108) | 2181 |
| Number of patients >100 Beats per min, n(%) | 857 (39) |  |
| Temperature in degree Fahrenheit | 99 (2) | 2180 |
| Number of patients >100.4-degree Fahrenheit, n (%) | 535 (25) |  |
| Respiratory Rate in breaths/minute | 20 (3) |  |
| Systolic Blood Pressure in mmHg, Median (IQR) | 130 (29) | 2176 |
| Diastolic Blood Pressure in mmHg, Median (IQR) | 74 (17) |  |
| Admission Laboratory Parameters, Median (IQR) |  |  |
| White Blood Cell Count -K/uL, Median (IQR) | 7 (5.2-9.9) | 2029 |
| Number of patients >10, n (%) | 498 (25) | 2029 |
| Number of patients <4, n (%) | 224 (11) | 2029 |
| Lymphocyte Count - K/uL, Median (IQR) | 0.9 (0.7-1.3) | 1888 |
| Lymphocyte Percentage, Median (IQR) | 13.8(8.5-20.5) | 1972 |
| Hemoglobin - g/dL, Median (IQR) | 13.1(11.6-14.4) | 2031 |
| Platelet Count - K/uL, Median (IQR) | 195(151-253) | 2028 |
| Prothrombin Time - sec, Median (IQR) | 13.9(13.2-15) | 937 |
| Activated Partial Thromboplastin Time - sec, Median (IQR) | 32 (29-36.1) | 926 |
| Serum Sodium - mEq/L, Median (IQR) | 137 (134-140) | 2015 |
| Serum Potassium - mEq/L, Median (IQR) | 4.2 (3.9-4.6) | 2006 |
| Serum Creatinine - mg/dL, Median (IQR) | 1 (0.8-1.5) | 2018 |
| Aspartate aminotransferase - U/L, Median (IQR) | 42 (30-70.5) | 1815 |
| Alanine aminotransferase - U/L, Median (IQR) | 32 (19-56) | 1025 |
| Serum Albumin - g/dL, Median (IQR) | 3.2 (2.8-3.6) | 1848 |
| Venous lactate - mmol/L, Median (IQR) | 1.5 (1.1-2) | 775 |
| Number of patients >1.5, n (%) | 392 (51) | 775 |
| Inflammatory Markers |  |  |
| C-Reactive Protein - mg/L, Median (IQR) | 110 (57.8-196) | 1354 |
| Ferritin -ng/L, Median (IQR) | 714 (324-1730) | 1324 |
| Procalcitonin -ng/mL, Median (IQR) | 0.18 (0.08-0.57) | 1241 |
| Number of patients >0.49, n (%) | 344 (28) | 1241 |
| Number of patients <0.15, n (%) | 573 (46) | 1241 |
| Lactate Dehydrogenase -U/L, Median (IQR) | 416 (314-545) | 1290 |
| Creatinine Kinase - U/L, Median (IQR) | 199 (97-565) | 502 |
| D-Dimer -ug/mL, Median (IQR) | 1.31 (0.74-2.44) | 1079 |
| Number of patients >2.0, n (%) | 352 (33) | 1079 |

Baseline values are defined as first values on hospitalization. All continuous characteristics are in Median (Interquartile Range) unless specified otherwise and all categorical characteristics are in number (percentage). The percentage is calculated with the number of patients who had the characteristic available as the denominator. For further clarity, the number in which that characteristic was available for is provided separately in adjacent column.

### Laboratory Results and Vital Signs at Presentation

Overall, 1,558 (71%) of patients were admitted through the emergency department. On hospital admission, 39% of all patients were tachycardic and 25% of all patients were febrile (**Table 1**). The median white blood cell count was 7 K/uL and lymphocyte percentage was 13.8. The median serum creatinine was 1 mg/dL. Select inflammatory markers performed in subsets of patients in accordance with clinical indication were markedly elevated on admission (**Table 1**). Specifically, the median C-reactive protein (CRP) was 110 mg/L, lactate dehydrogenase (LDH) was 416 U/L, and ferritin was 714 ng/L. Over one-quarter of patients (28%) had a procalcitonin level above 0.49 ng/mL, and nearly half of patients (46%) had a procalcitonin level less than 0.15 ng/mL. The median D-dimer was 1.31 ug/mL; one-third of patients (33%) had a D-dimer greater than 2 ug/mL.

The frequencies of otherwise non-routine laboratory assessments ordered on day of admission increased over time and are shown in **Supplementary Figure 2**. In contrast, hemoglobin, a routinely measured clinical lab value, was ordered at admission in the majority of patients without variation over the study period.

### Clinical Outcomes

Due to the unknown future clinical course of those patients hospitalized at the time of data freeze, below we present clinical characteristics of only those patients who had completed their hospital course. A total of 1,078 Covid-19 confirmed hospitalized patients completed their hospital course (died or discharged alive) by the date of data freeze on April 02, 2020. Of these, 768 (71%) were discharged and 310 (29%) died in the hospital. Estimates for mortality and need for ICU admissions over time are displayed in **Figure 2**.

**Figure 2:**
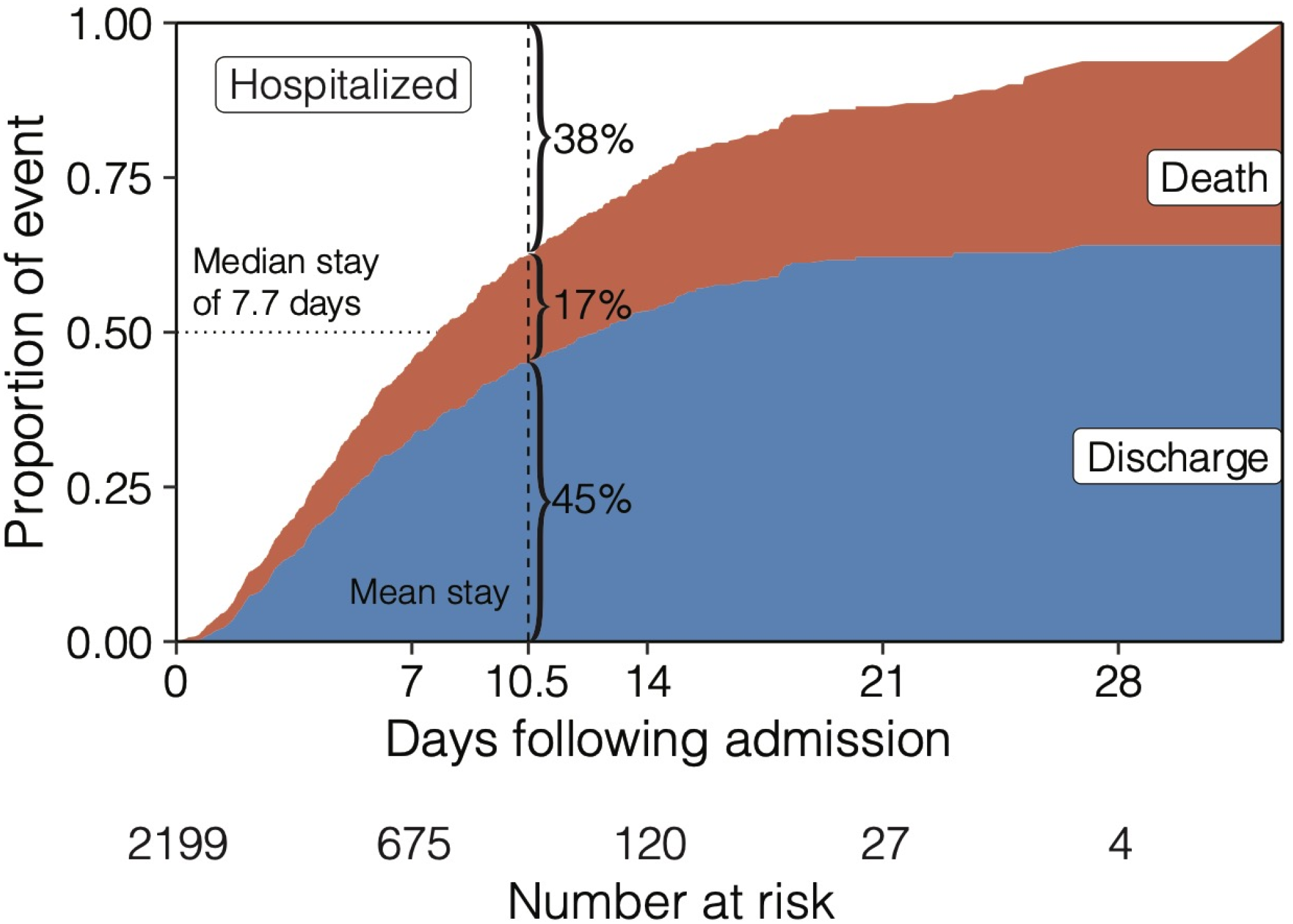
Cumulative incidence function displays the probability of mutually exclusive events of discharge (blue) or death (red) by a given day of hospitalization, accounting for the changing number of patients at risk including censoring. The remaining portion (white) shows patients that are still hospitalized, where the median length of stay is 7.7 days.

The median LOS was 7.7 days, accounting for censoring of patients with ongoing hospitalization. By the mean LOS (10.5 days), 45% of patients had been discharged, 17% had died, and 38% were still hospitalized (**Figure 2**). Demographics and admission laboratory measurements for patients who completed their hospital course are displayed in **Table 2**, stratified by mortality. The median age was 75 years in those who died and 59 in those who were discharged. Pre-existing conditions were present in 64% of those who died and 46% of those discharged. Among those who died, 45% had hypertension, 34% had type 2 diabetes, 27% had coronary artery disease, 21% had heart failure and 13% had chronic kidney disease. In patients who were discharged, 30% had hypertension, 20% had type 2 diabetes, 11% had coronary artery disease, 7% had heart failure and 7% had chronic kidney disease.

**Table 2.** Characteristics of Hospitalized Covid-19 patients by patients who had in-hospital mortality vs. those who were discharged alive (N=1,078)

|  | Patients with In-Hospital Mortality (n=310) | N with characteristic available | Patients who were discharged alive (n=768) | N with characteristic available |
| --- | --- | --- | --- | --- |
| Admitted to Intensive Care, n (%) | 121 (39) | 310 | 264 (34.4) | 768 |
| Race, n (%) |  |  |  |  |
| White | 98 (31.6) | 310 | 216 (28.1) | 768 |
| Black or African American | 71 (22.9) |  | 185 (24.1) |  |
| Asian | 17 (5.5) |  | 22 (2.9) |  |
| Pacific Islander | 1 (0.3) |  | 7 (0.9) |  |
| Other | 111 (35.8) |  | 310 (40.4) |  |
| Unknown | 12 (3.9) |  | 28 (3.6) |  |
| Ethnicity, n (%) |  |  |  |  |
| Hispanic/Latino | 58 (18.7) | 310 | 208 (27.1) | 768 |
| Non-Hispanic/Latino | 198 (63.9) |  | 464 (60.4) |  |
| Unknown | 54 (17.4) |  | 96 (12.5) |  |
| Age, Median (IQR) | 75 (64-85) |  | 59 (45-72) |  |
| Age Groups, n (%) |  |  |  |  |
| 18-30 | 1 (0.32) | 310 | 53 (6.9) | 768 |
| 31-40 | 2 (0.65) |  | 108 (14.1) |  |
| 41-50 | 17 (5.5) |  | 100 (13.0) |  |
| 51-60 | 37 (11.9) |  | 143 (18.6) |  |
| 61-70 | 66 (21.3) |  | 157 (20.4) |  |
| 71-80 | 83 (26.8) |  | 130 (16.9) |  |
| 80-90 | 75 (24.2) |  | 62 (8.1) |  |
| >90 | 29 (9.4) |  | 15 (2.0) |  |
| Sex, n (%) |  |  |  |  |
| Male | 191 (61.6) | 310 | 436 (56.8) | 768 |
| Female | 119 (38.4) |  | 332 (43.2) |  |
| Body Mass Index in kg/m<sup>2</sup>, Median (IQR) | 32 (7) | 241 | 28 (6) | 627 |
| Previous Medical History, n (%) |  |  |  |  |
| Atrial Fibrillation | 43 (13.9) | 768 | 42 (5.5) | 768 |
| Asthma | 23 (7.4) |  | 61 (7.9) |  |
| Coronary Artery Disease | 83 (26.8) |  | 84 (10.9) |  |
| Cancer | 24 (7.7) |  | 40 (5.2) |  |
| Chronic Kidney Disease | 41 (13.2) |  | 54 (7) |  |
| Chronic Obstructive Pulmonary Disease | 28 (9) |  | 31 (4) |  |
| Diabetes Mellitus | 105 (33.9) |  | 151 (19.7) |  |
| Heart Failure | 64 (20.6) |  | 53 (6.9) |  |
| Hypertension | 140 (45.2) |  | 233 (30.3) |  |
| Stroke | 32 (10.3) |  | 40 (5.2) |  |
| **Admission Laboratory Parameters** |  |  |  |  |
| White Blood Cell Count -K/uL, Median (IQR) | 8.6 (5.9-12) | 281 | 6.2 (4.7-8.2) | 728 |
| Number of patients >10, n (%) | 105(37) | 281 | 101(14) | 728 |
| Number of patients <4, n (%) | 17 (6) | 281 | 106 (15) | 728 |
| Hemoglobin - g/dL, Median (IQR) | 12.6 (10.9-14.3) | 282 | 13.2 (11.9-14.4) | 728 |
| Platelet Count - K/uL, Median (IQR) | 185 (144-239) | 282 | 193 (153-247) | 726 |
| Lymphocyte Count - K/uL, Median (IQR) | 0.9 (0.6-1.3) | 248 | 1 (0.8-1.4) | 689 |
| Lymphocyte Percentage, Median (IQR) | 11.7 (6.6-18.6) | 270 | 16.6 (11.7-24.2) | 710 |
| Prothrombin Time - sec, Median (IQR) | 14.2 (13.6-16.3) | 142 | 13.5 (13-14.4) | 304 |
| Activated Partial Thromboplastin Time - sec, Median (IQR) | 32.9 (30.2-37.6) | 140 | 31.4 (28.7-34.8) | 302 |
| Serum Sodium - mEq/L, Median (IQR) | 138 (134-141) | 284 | 137 (135-140) | 708 |
| Serum Potassium - mEq/L, Median (IQR) | 4.4 (4-5) | 288 | 4.1 (3.8-4.5) | 706 |
| Serum Creatinine - mg/dL, Median (IQR) | 1.3 (0.9-2.2) | 288 | 0.9(0.7-1.2) | 710 |
| Aspartate aminotransferase - U/L, Median (IQR) | 58.5 (34-102) | 246 | 36 (26-54) | 628 |
| Alanine aminotransferase - U/L, Median (IQR) | 32 (19-58) | 119 | 30 (19-52) | 358 |
| Serum Albumin - g/dL,<br>Median (IQR) | 3 (2.7-3.4) | 255 | 3.4 (3-3.7) | 635 |
| Venous lactate -<br>mmol/L, Median (IQR) | 1.8 (1.3-2.6) | 108 | 1.3 (1.1-1.8) | 236 |
| Number of patients<br>>1.5 mmol/L, n (%) | 74 (69) | 108 | 88 (37) | 236 |
| **Inflammatory Markers** |  |  |  |  |
| C-Reactive Protein -<br>mg/L, Median (IQR) | 162 (75.7-266) | 141 | 79.3 (37.5-144) | 422 |
| Ferritin -ng/L, Median<br>(IQR) | 798 (397-2020) | 146 | 509 (235-987) | 410 |
| Procalcitonin -ng/mL,<br>Median (IQR) | 0.44 (0.16-1.45) | 138 | 0.09 (0.05-0.22) | 392 |
| Number of patients<br>>0.49, n (%) | 67 (49) | 138 | 46 (12) | 392 |
| Number of patients<br><0.15, n (%) | 34 (25) | 138 | 263 (67) | 392 |
| Lactate<br>Dehydrogenase -U/L,<br>Median (IQR) | 517(371-756) | 134 | 347 (272-448) | 419 |
| Creatinine Kinase -<br>U/L, Median (IQR) | 472 (139-940) | 74 | 154 (84-319) | 126 |
| D-Dimer -ug/mL,<br>Median (IQR) | 2.41 (1.18-3.79) | 117 | 0.93 (0.58-1.61) | 282 |
| Number of patients<br>>2.0, n (%) | 66(56) | 117 | 59 (21) | 282 |

Baseline values are defined as first values on hospitalization. All continuous characteristics are in Median (Interquartile Range) unless specified otherwise and all categorical characteristics are in number (percentage). The percentage is calculated with the number of patients who had the characteristic available as the denominator. For further clarity, the number in which that characteristic was available for is provided separately in adjacent column.

We present key laboratory markers at the time of hospital admission in subsets of patients for whom they were measured. In patients who died, the median lymphocyte percentage was 11.7%, aspartate aminotransferase (AST) was 58.5 U/L, CRP was 162 mg/L, D-dimer was 2.41 ug/mL (56% had a D-dimer >2 ug/mL), ferritin was 798 ng/L, LDH was 517 U/L, creatinine kinase (CK) was 472 U/L and procalcitonin was 0.44 ng/mL (49% had a procalcitonin >0.49 ng/mL). In patients who were discharged, the median lymphocyte percentage was 16.6%, AST was 36 U/L, CRP was 79.3 mg/L, D-dimer was 0.93 ug/mL (21% had a D-dimer >2 ug/mL), ferritin was 509 ng/L, LDH was 347 U/L, CK was 154 U/L and procalcitonin was 0.09 ng/mL (12% had a procalcitonin >0.49 ng/mL).

### Patients Requiring ICU Admission

Of the 1,078 patients who completed their hospital course, 385 (36%) required intensive care during their hospital stay. For these patients, vital signs and laboratory values immediately before transfer to intensive care are displayed in **Table 3**, stratified by mortality outcome. Among patients who died, 38% were tachycardic and 22% were hypotensive. Among patients who were eventually discharged, 19% were tachycardic and 4% were hypotensive. In patients who died, the median lymphocyte percentage was 9.6%, serum creatinine was 1.5 mg/dL, AST was 62 U/L, CRP was 220 mg/L, ferritin 920 ng/L, procalcitonin was 1.02 ng/mL (59% had a procalcitonin >0.49 ng/mL), LDH was 513 U/L, CK was 659 U/L and D-dimer was 2.7 ug/mL (63% had a D-dimer >2 ug/mL). Among patients who were discharged, the median lymphocyte percentage was 16.6%, serum creatinine was 0.8 mg/dL, AST was 35 U/L, CRP was 75.6 mg/L, ferritin 503 ng/L, procalcitonin was 0.08 ng/mL (10% had a procalcitonin >0.49 ng/mL), LDH was 333 U/L, CK was 146 U/L and D-dimer was 0.83 ug/mL (22% had a D-dimer >2 ug/mL).

**Table 3.**
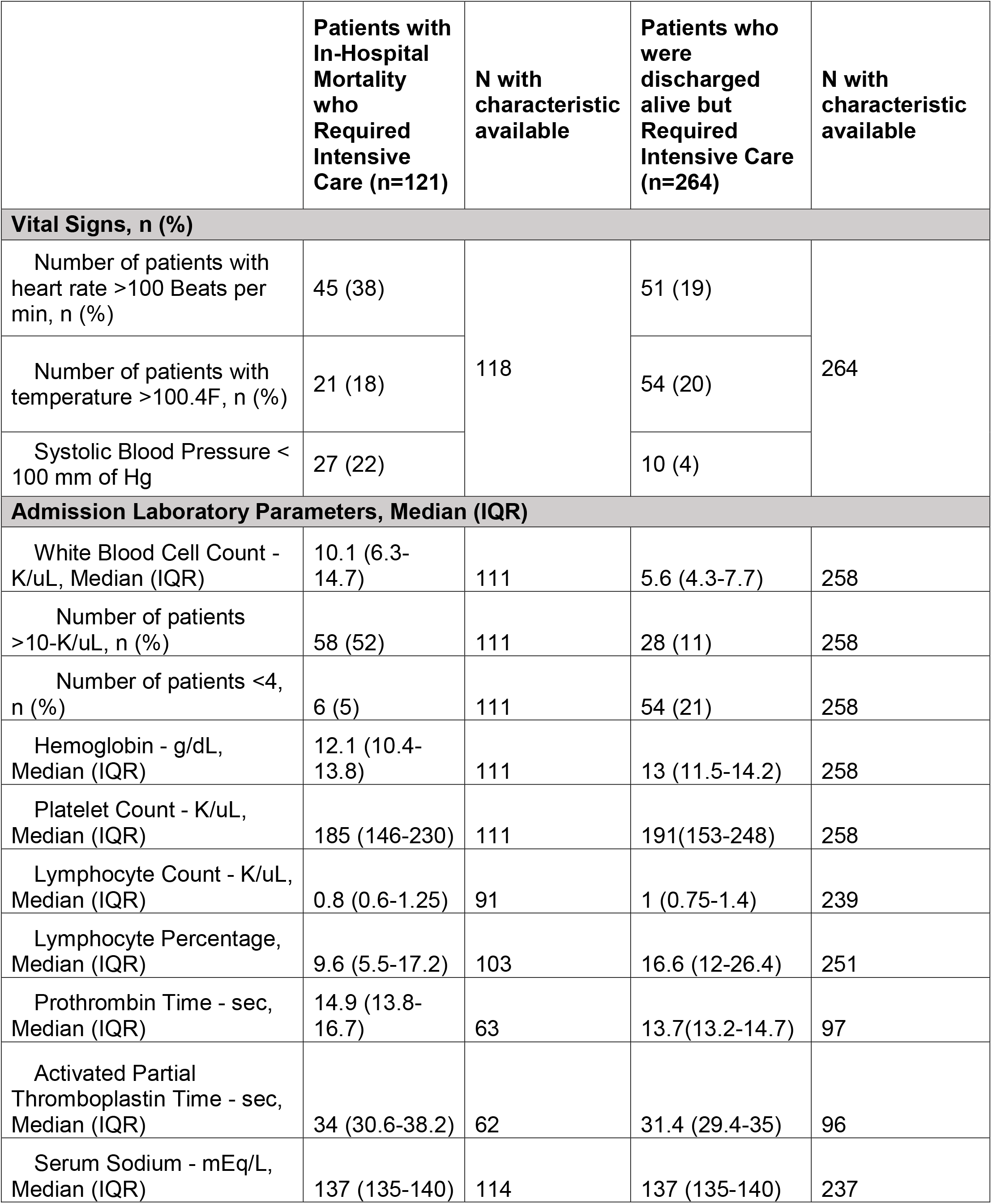
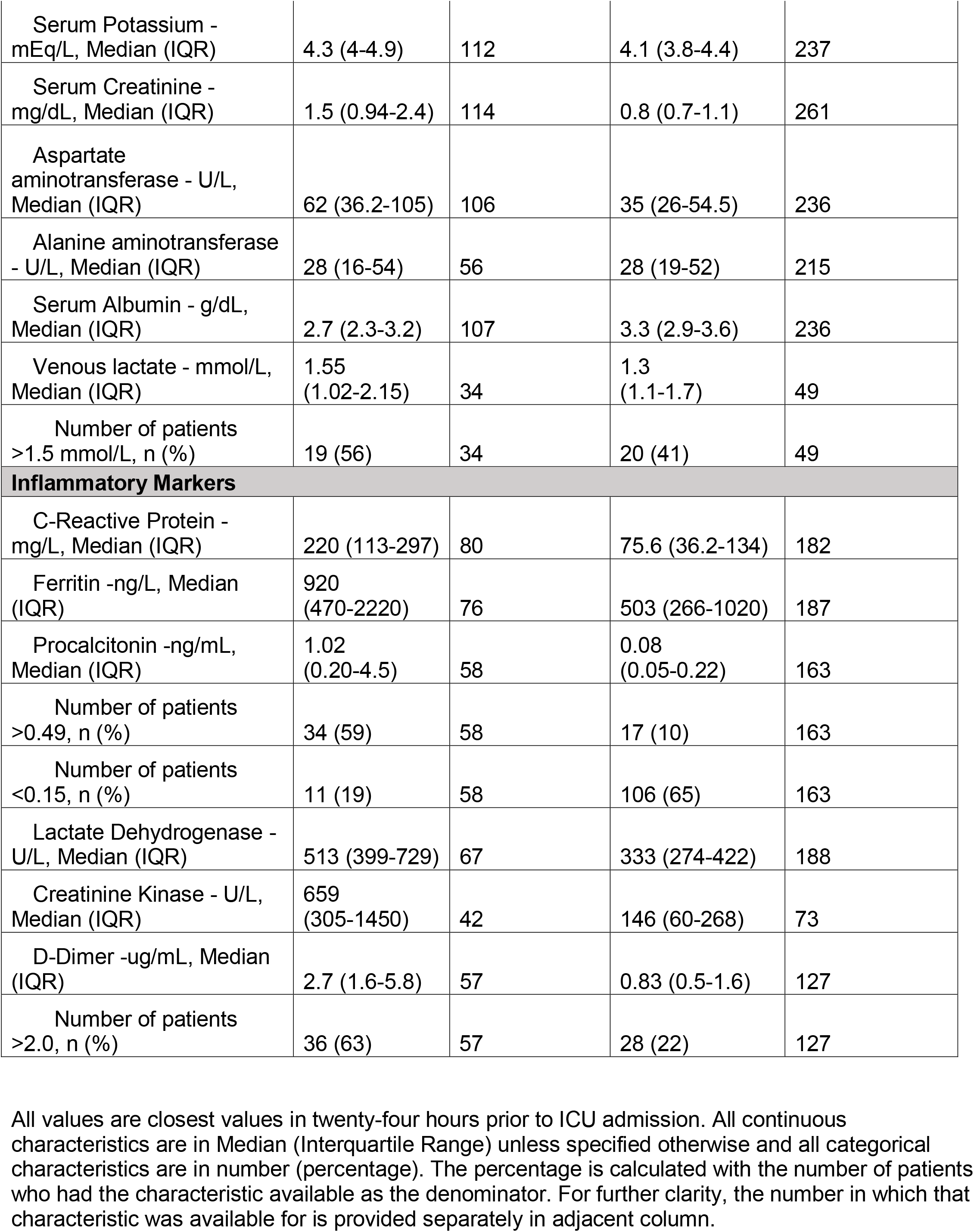
Selected characteristics for hospitalized Covid19 patients before transfer to Intensive care stratified by in-hospital mortality (N= 385)

## DISCUSSION

The Covid-19 pandemic represents the greatest public health emergency in the modern world. Limited data, especially in the US, exists to guide clinical care, resource management, and risk stratification in hospitalized patients. To the best of our knowledge, our study is the largest and most diverse case series of patients reported with confirmed Covid-19 in the US. Previous reports were either from other countries, examined smaller cohorts, or were focused on critically ill patients^4,8–13^ The present report provides a broad perspective on patients admitted with Covid-19 in both general medicine ward and intensive care settings. Additionally, our health system serves a unique population representative of the ethnic and socioeconomic diversity seen in both New York City and across the United States.

We highlight several key findings. Among the 1,078 patients who completed their hospital course (discharge or in-hospital death), the overall mortality rate was 29% and 31% in patients who received ICU care. The overall case fatality rate likely represents an overestimation of the true disease mortality rate since patients who remained hospitalized at the date of data freeze were not included in this calculation. The mortality rate in intensive care is lower than previously described^4,13–15^. and may be reflective of early care escalation or the larger number of patients in our study.

Though no formal comparisons were made, we observed that patients who died had a higher median age and a greater frequency of male sex with more pre-existing conditions than those who were discharged. Though 25% of patients were febrile on admission, this may be an underestimation due to possible antipyretic use and/or selection bias. A substantial proportion of patients with Covid-19 displayed abnormal laboratory measurements at the time of admission. These included lymphopenia and elevated inflammatory markers such as D-dimer, CRP, LDH, and ferritin. These trends persisted among those who died and/or received intensive care, both on admission and at the time of ICU transfer. If formal epidemiologic analyses confirm these observations, early laboratory evaluation may be crucial in identifying patients suspected for Covid-19 prior to RT-PCR test result. It may also aid clinicians in identifying patients at high risk of decompensation, ICU admission, and potentially even death. Early identification of high-risk patients could enable timely patient triage and improved resource allocation. Additional work is needed to develop real-time, accurate predictive models for risk stratification in Covid-19, particularly to elucidate the clinical utility of specific laboratory measurements.

Han et al. found a significant association of serum lactate dehydrogenase and c-reactive protein with Covid-19 severity in patients from China^16^. As the number of cases rose quickly in New York City, MSHS hospitals served as early adopters, creating a Covid-19 order set in our EHR to streamline objective data gathering, facilitate more cohesive workflow amongst team members, and minimize ancillary staff exposure by completing all necessary admission labs at one time. This laboratory order set included serum D-dimer, C-reactive protein, procalcitonin, ferritin, and lactate dehydrogenase. In turn, we observed an increase in these orders from the first day of admission over the study period (**Supplementary Figure 2**). Given the abnormalities observed in patients who died, these laboratory measurements may be prognostic markers of disease severity or subsequent clinical course, although this requires further investigation. If confirmed, other health systems expecting impending case surges may consider similar workflows to promote improved healthcare delivery to affected patients.

Our study should be considered in light of several limitations. Since Covid-19 testing is frequently repeated in hospitalized patients and initial testing may result in false negatives, we are unable to determine whether patients developed their infection during or before hospital admission. Furthermore, Covid-19 has a variable incubation period of approximately 8-15 days^17^, and patients may present to the hospital several days after initial infection or the onset of symptoms. Thus, we are unable to determine patients’ disease duration. Additionally, we separated discharged patients from those who died, but some patients may have expired after discharge. This could affect our listed case mortality rate. Our study is also confined by the inherent limitations (e.g. biases) of EHR data. Although utilizing structured EHR data allows for rapid integration of multiple data streams and real-time analysis, data present only in clinical note text, such as symptoms on presentation are missed. We chose not to perform comprehensive manual chart review to prioritize timely dissemination of our observations.

As the Covid-19 pandemic spreads from the current epicenter in New York City to other areas, our report provides meaningful clinical insights that may better inform care for diverse populations. Future work will aim to predict Covid-19 patient outcomes using a variety of approaches, thereby reducing healthcare system burden and permitting improved care delivery.

## Data Availability

NA

